# ChatGPT in medical literature – a concise review and SWOT analysis

**DOI:** 10.1101/2023.05.06.23289608

**Authors:** Daniel Gödde, Sophia Nöhl, Carina Wolf, Yannik Rupert, Lukas Rimkus, Jan Ehlers, Frank Breuckmann, Timur Sellmann

## Abstract

**Background:** ChatGPT (Chat Generative Pre-trained Transformer) has initiated widespread conversation across various human sciences. We here performed a concise review combined with a SWOT (strengths, weaknesses, opportunities, threats) analysis on ChatGPT potentials in natural science including medicine.

**Methods:** This is a concise review of literature published in PUBMED from 01.12.2022 to 31.03.2023. The only search term used was “ChatGPT”. Publications metrics (author, journal, and subdisciplines thereof) as well as findings of the SWOT analysis are presented.

**Findings:** Of 178 studies in total, 160 could be evaluated. The average impact factor was 4,423 (0 – 96,216), average publication speed was 16 days (0-83 days). Of all articles, there were 77 editorials, 43 essays, 21 studies, six reviews, six case reports, six news, and one meta-analyses. Strengths of ChatGPT include well-formulated expression as well as the ability to formulate general contexts flawlessly and comprehensibly, whereas the time-limited scope as well as the need for correction by experts were identified as weaknesses and threats. Opportunities include assistance in formulating medical issues for non-native speakers as well as the chance to be involved in the development of such AI in a timely manner.

**Interpretation:** Artificial intelligences such as ChatGPT will revolutionize more than just the medical publishing landscape. One of the biggest dangers in this is uncontrolled use, so we would do well to establish control and security measures at an early stage.

**Research in context:** *Evidence before this study:* Since its release in 11/ 2022, only a few randomized controlled trials using ChatGPT have been published. To date, the majority of data stems from short notes or communication. Given the enormous interest (and also potential for misuse), we conducted a PUBMED literature search to create the most comprehensive evidence base currently available. We searched PUBMED for publications including the quote “ChatGPT” in English or German from 01.12.2022 until 31.03.2023. In order not risk any bias of evidence all related publications were screened initially.

*Added value of this study:* This is the most concise review for ChatGPT up to date. By means of a SWOT analysis, readers and researchers gain comprehensive insight to strengths, weaknesses, opportunities and threats of ChatGPT especially in the context of medical literature.

*Implications of all the available evidence:* Our review may well serve as origin for further research related to the topic in order to create more evidence, strict regulations and policies in dealing with ChatGPT.

## Introduction

Chat Generative Pre-trained Transformer (ChatGPT), is a 175-billion-parameter natural language processing model that can generate conversation-style responses to user input. Since its launch by OpenAI, ChatGPT has stimulated widespread conversation and momentum across various human sciences, including medicine. It enables humans to interact and discuss with an artificial intelligence chatbot on a broad range of topics. At the latest since it became known that ChatGPT was able to pass the United States Medical Licensing Examination performing at a level comparable to a third-year medical student,^1^ it moved into the center of interest. Preliminary evidence exists, that ChatGPT has promising applications across the clinical workflow.^2^ Moreover, ChatGPT and similar generative AI have specific applications within medical education, including clinical vignette generation and communications training with AI virtual patients;^2–4^ however, it also poses several challenges that universities need to carefully address.^5^ Tested on the verbal scale of the Wechsler adult intelligent scale (WAIS), an intelligence quotient of 155 was demonstrated for ChatGPT.^6^ When asked, ChatGPT itself explains its role in medical literature in the following way: “*As an AI language model, ChatGPT’s role in medical literature is to provide information and insights related to healthcare and medicine based on its knowledge of the subject matter. ChatGPT can help researchers and healthcare professionals by providing quick access to information on a wide range of medical topics, including symptoms, diagnoses, treatments, and outcomes. ChatGPT can also assist in generating summaries and abstracts of medical articles and papers, making it easier for healthcare professionals to keep up with the latest developments in their field. Additionally, ChatGPT can assist in developing natural language processing applications that can improve the efficiency and accuracy of medical data analysis and interpretation. However, it’s important to note that ChatGPT is not a substitute for medical professionals and should not be used to make medical decisions. It is always recommended to consult with a licensed healthcare provider for medical advice and treatment*.”

Although these functions are promising and tempting, the technology is still in its early stages, making the program vulnerable to mis- and abuse. Since its release, over 200 papers have addressed issues using the search term “ChatGPT”. The aim of this review was to analyze the role ChatGPT in medical literature during the first three months after its release and put its use in various contexts. In order to provide an overview and to find out more about its potential as well as to create hypotheses for further research, we undertook this concise review on the role of ChatGPT including a SWOT (strengths, weaknesses, opportunities and threats) analysis to define its potential especially for medical literature.

## Methods

### Study design

#### Search strategy and selection criteria

References for this Review were identified through searches of PubMed with the search term “ChatGPT” from 01-12-2022 until 31-03-2023. Only fully retrievable papers published in English or German and were reviewed. The final reference list was generated on the basis of originality and relevance to the broad scope of this Review.

### Data analysis

Only complete data sets published in English or German with respect to the above criteria were included in this study. Additional exclusion criteria included incomplete or non-retrievable data sets as well as articles completely written *completely by* but not *about* ChatGPT. All accessible publications were evaluated according to the following specifications by the author team.

#### Articles

Articles were primarily classified according to the specifications of PUBMED. For better comprehensibility, a “studies” category was created, defined as “a method of research in which a problem is identified, relevant data are gathered, a hypothesis is formulated, and the hypothesis is empirically tested”. All identified articles were scanned for “qualitative” (collection of text-based data, e.g. interviews, focus groups, usually hypothesis generating) vs. “quantitative” (collection of number-based data, e.g. measurements, questionnaires with associated statistics, usually hypothesis-testing) content. We also chose to discriminate “mixed method research” (combination of qualitative and quantitative content) and “reviews and meta-analyses”. Furthermore, empirical data, based on a (proprietary) database, was distinguished from non-empirical data (including anything without a database). Article content was analyzed in reporting on the use or actual, partial or full use of ChatGPT in the drafting of the article. In this context, attention was also paid to the correlation between the share of ChatGPT in the preparation of the manuscript (not at all, partially, completely) and the achievable impact factor. In order to better compare the course of the number of actual published papers, an article count was displayed by week and compared to weekly article release during the COVID-19-outbreak.

#### Journals

Journals publishing articles on ChatGPT were evaluated regarding title, discipline of natural science, actual impact factor, open access vs. traditional publishing and publishing speed (incl. preprint server).

#### Authors

To obtain more information about authors publishing on the topic, the number of first and last authorships other than the index publication was determined for the years 2020 through 2022. Additionally, the specialty, if identifiable via PUBMED or the full text (ORCID ID), was also reported.

#### SWOT analysis

During the screening of all evaluated articles, quotes on strengths, weaknesses, opportunities and threats mentioned within the articles were collected. Subsequently, the items identified were evaluated in a Delphi round, consented upon and assigned in keyword form to one of the four components of the SWOT analysis.

### Statistical analysis

The primary endpoint was applicable extent of data collection on the role of ChatGPT in medical literature defined by author, article and journal type. Secondary endpoints included strengths, weaknesses, opportunities and threats of Chat GPT use in medical literature. All data were analyzed on a descriptive basis. Data are means ± SD unless otherwise stated. Statistical analysis was performed using descriptively using Microsoft Excel for Office 365 (Version16.16.27) and PSPP, Version 1.6.2 (https://www.gnu.org/software/pspp/). Student’s t-test, Levene’s test, and Mann-Whitney-U test were applied as appropriate. A *p* < 0.05 was considered to represent statistical significance.

## Results

From 01.12.2022 until 31.03.2023, a total of 178 papers using the search term “ChatGPT” were published in PUBMED, thereof six papers in December 2022, sixteen in January, 68 in February and 88 in March 2023. After a thorough human review, eighteen papers had to be excluded, 11 papers because they were written *with* but not *about* ChatGPT, four papers were not retrievable as full text, and two papers were neither written in English nor German. One paper was just an erratum note. Figure 1 shows the PRISMA flow chart of ChatGPT related publications.

**Figure 1.**
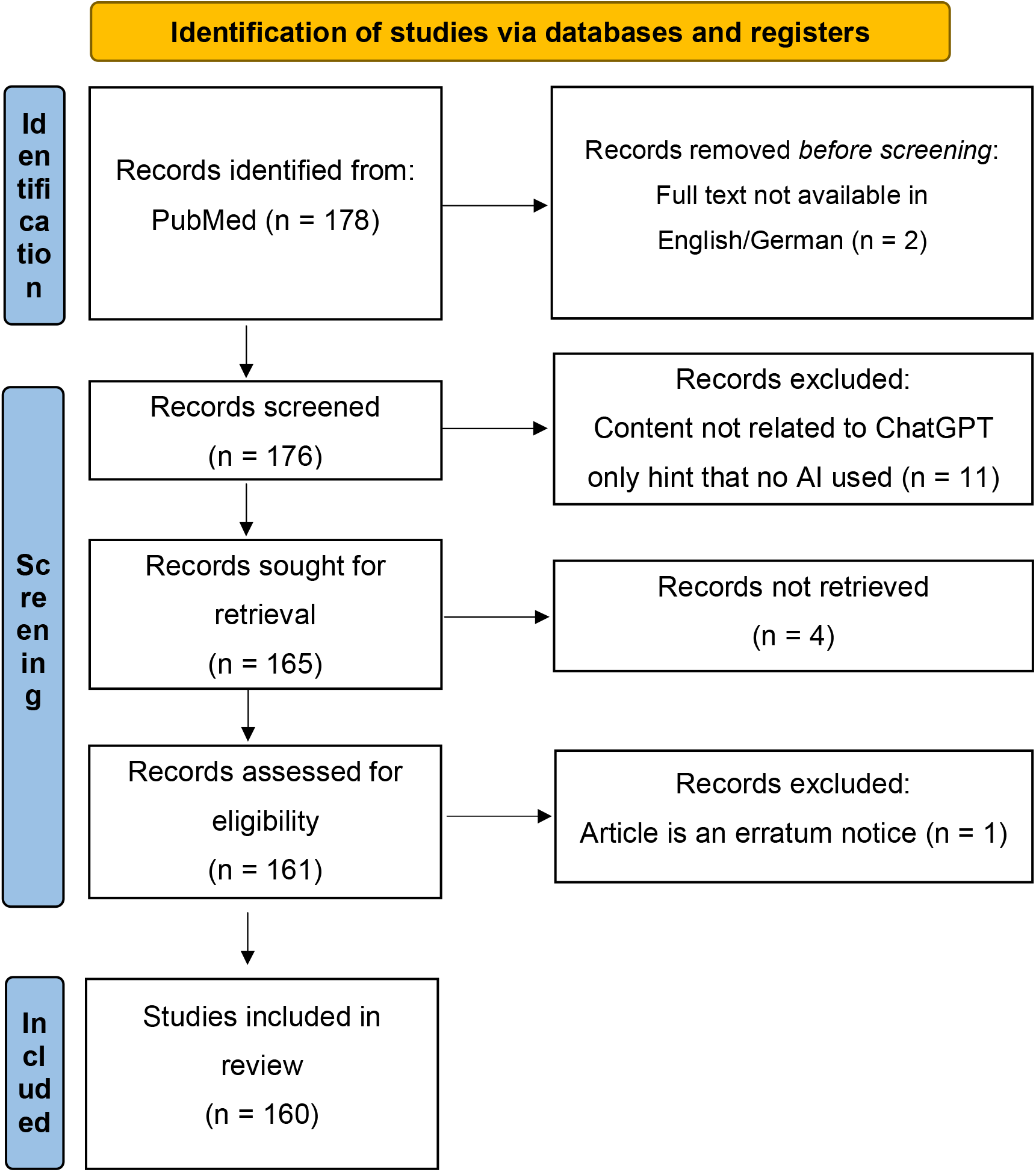
PRISMA flow chart of ChatGPT related studies (modified after ^7^)

### Articles

There vast majority of all articles were brief statements like editorials, or letters to the editor (48·1%, n = 77). Essays or commentaries (26·9%, n = 43) represented the second largest portion of the articles. Studies (13·3%, n = 21), reviews, news, and case reports (each 3·8%, each n = 6), or meta-analysis (0·6%, n = 1) were less frequent. No randomized controlled trial could be identified. Figure 2 shows the distribution of article types according to the specifications of PUBMED. Of all articles, 80% (n = 128) contained non-empirical and 20% (n = 32) contained empirical data. Of these again, 6·9% (n = 11) were of qualitative, 8·8% (n = 14) were of quantitative, and 1·9% (n = 3) of mixed nature. Regarding the proportion of ChatGPT within the article, 11·9% (19) of all articles were written at least partially with ChatGPT. The average impact factors are displayed in table 1.

**Figure 2.**
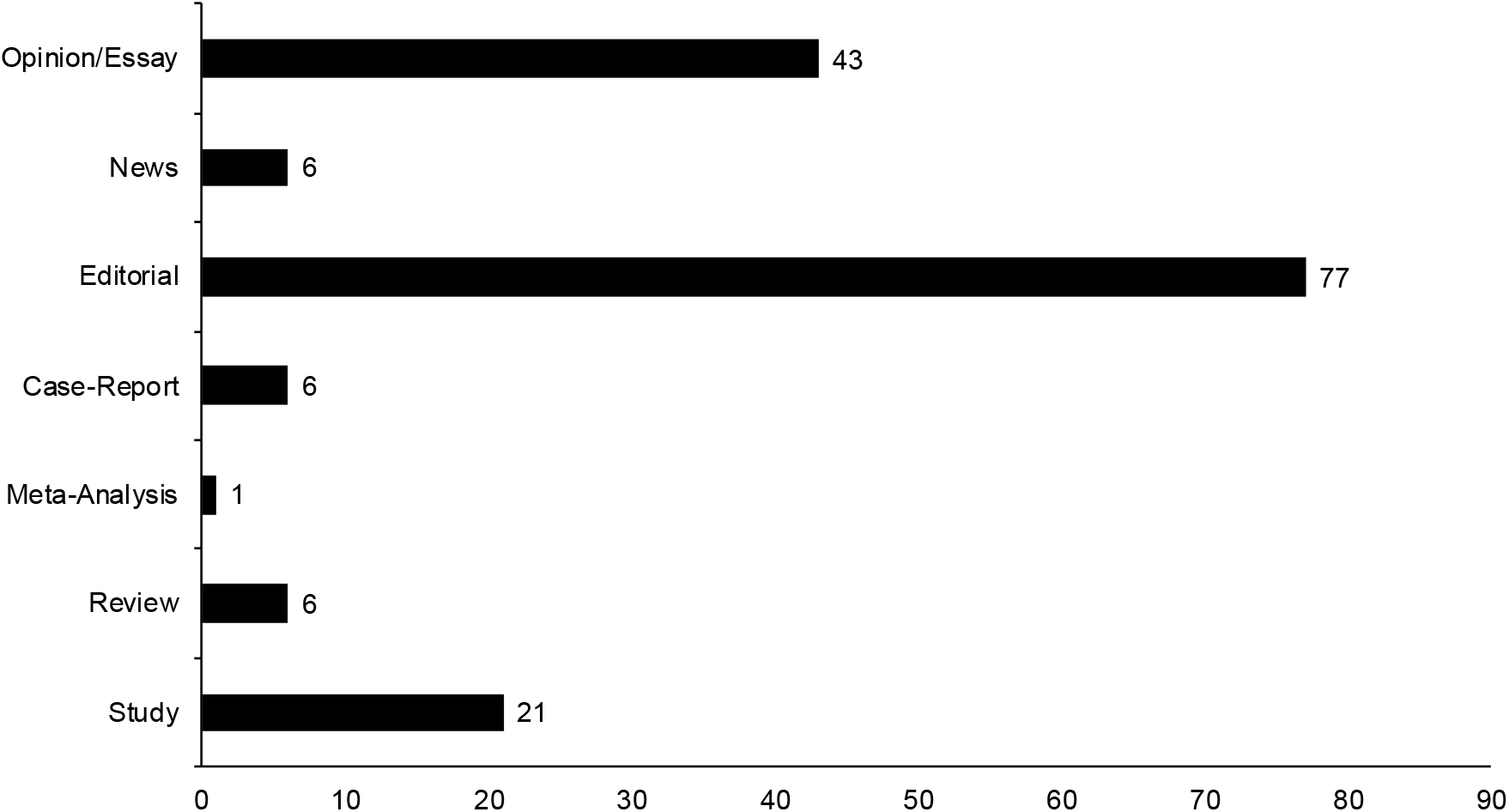
Distribution of article types regarding to PUBMED classification.

**Table 1.** Average impact for articles written with or without ChatGPT.

| Articles with impact factor written ... |  |  |  |  |
| --- | --- | --- | --- | --- |
|  | n | median | IQR | range |
| ... without ChatGPT | 120 | 5.622* | 3.282 – 13.89 | 0.646 – 96.216 |
| ... with ChatGPT | 18 | 4.403* | 3.499 – 7.188 | 1.15 – 29.983 |
| *$p$ = not significant | | | | |

\*Levene's test ( $p = 0.001$ ) indicated that the T-test, although primarily significant ( $p = 0.003$ ), was not robust, so statistical significance was again calculated using the Mann-Whitney U-test ( $p = 0.4$ ).

In order to illustrate scientific interest in the topic, as measured by number of publications, figure 3 shows the comparison to the number of Covid-19 publications during the first 12 weeks in 2020.

**Figure 3.**
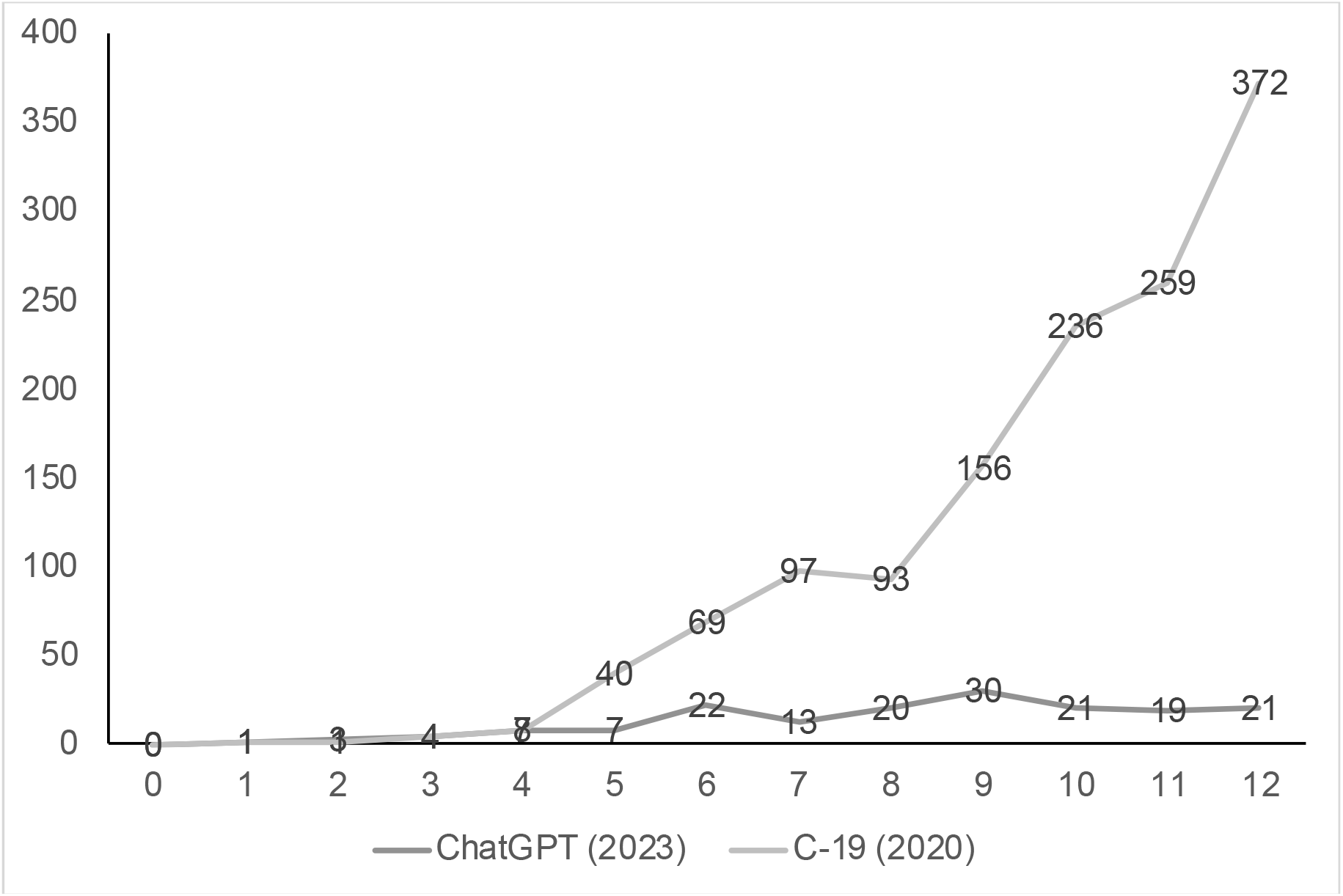
Weekly publications on ChatGPT (2023) compared to COVID-19 (2020) Data for C-19 related publications were taken from Kambhampati et al.^8^ During the first four weeks, there was no marked differences in publications (Chat GPT/C-19, week 1: 1/1, week 2: 3/1; week 3: 4/4; week 4: 7/8).

### Journals

Publishing journals showed a wide range of scientific disciplines. Figure 4a shows an overview of the specialty distribution of the journals. The current impact factor of the represented journals ranged from 0 to 96·216 with a median of 5·144 (IQR 3·352-11·325). Overall, 45·6% (73) of all articles were published “traditionally” in contrast 54·4% (87) that were published as “open access”. Of those two groups, 5% of “traditional” and 11% of “open access” publications were provided on preprint servers in advance. Data on publication speed was accessible in 33·1% of all evaluated articles. The average time to publication was 16 days, ranging from 4 to 83 days.

**Figure 4.**
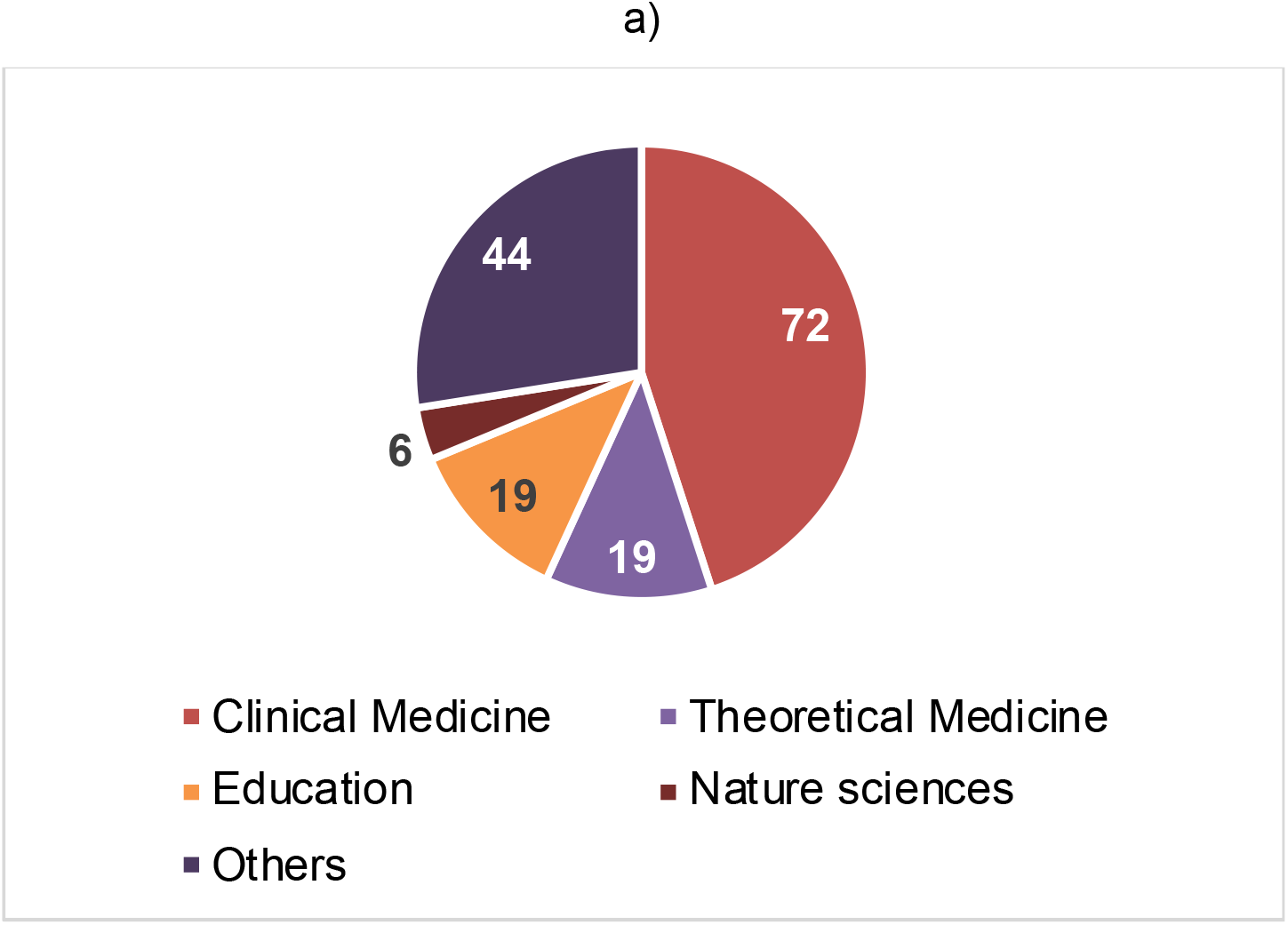
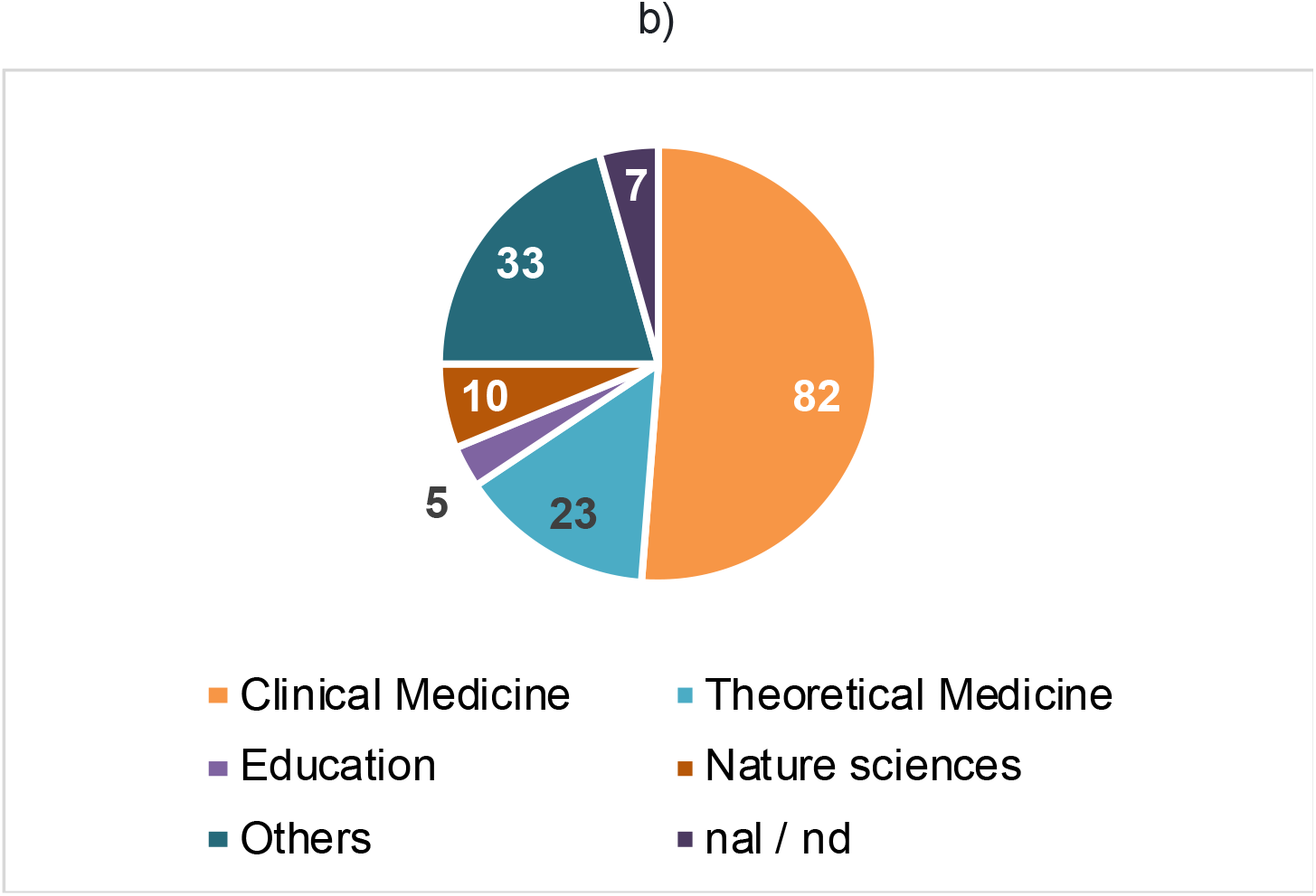
Disciplines of journals (a; up) and authors (b; down) publishing on ChatGPT. Clinical medicine includes all areas with direct patient contact (i.e. surgery or internal medicine), theoretical medicine areas without (i. e. microbiology or pharmacology). nal = no author listed; nd = not determinable

### Authors

Authors had a median of five (IQR 1-12 / range 0-94) first and a median of one last authorships (IQR 0-6 / range 0-61) in the years 2020-2022. Their area of expertise spanned all medical specialties up to science journalism, bioinformatics, nursing as well as humanities, economics and law. Figure 4 gives an overview of the specialty distribution of first authors (4b) publishing on ChatGPT.

### SWOT analysis

We were able to detect over 400 quotes in which information was provided on strengths, weaknesses, opportunities and threats. Of those, by far the most were related to weaknesses and least to opportunities. Quotes on strengths and threats were mentioned less frequently. Among the most prevalently cited weaknesses were limited abilities,^9,10^ lack of accuracy/ correctness,^11,12^ citation problems,^13,14^ and need for verification.^15,16^ Strengths, on the other hand, included reduced workload,^11,17–19^ data summarization,^20^ and high-quality results.^12,21–25^ Amongst the threats captured most frequently were plagiarism/ hallucination, scientific misconduct and ethical concerns,^22,26–28^ whereas major opportunities were seen in supporting different faculties.^21,29,30^ Due to the variability in the mentions, we decided to use a semiqualitative analysis. Results, conclusions and suggestions can be found in Figure 5.

**Figure 5.**
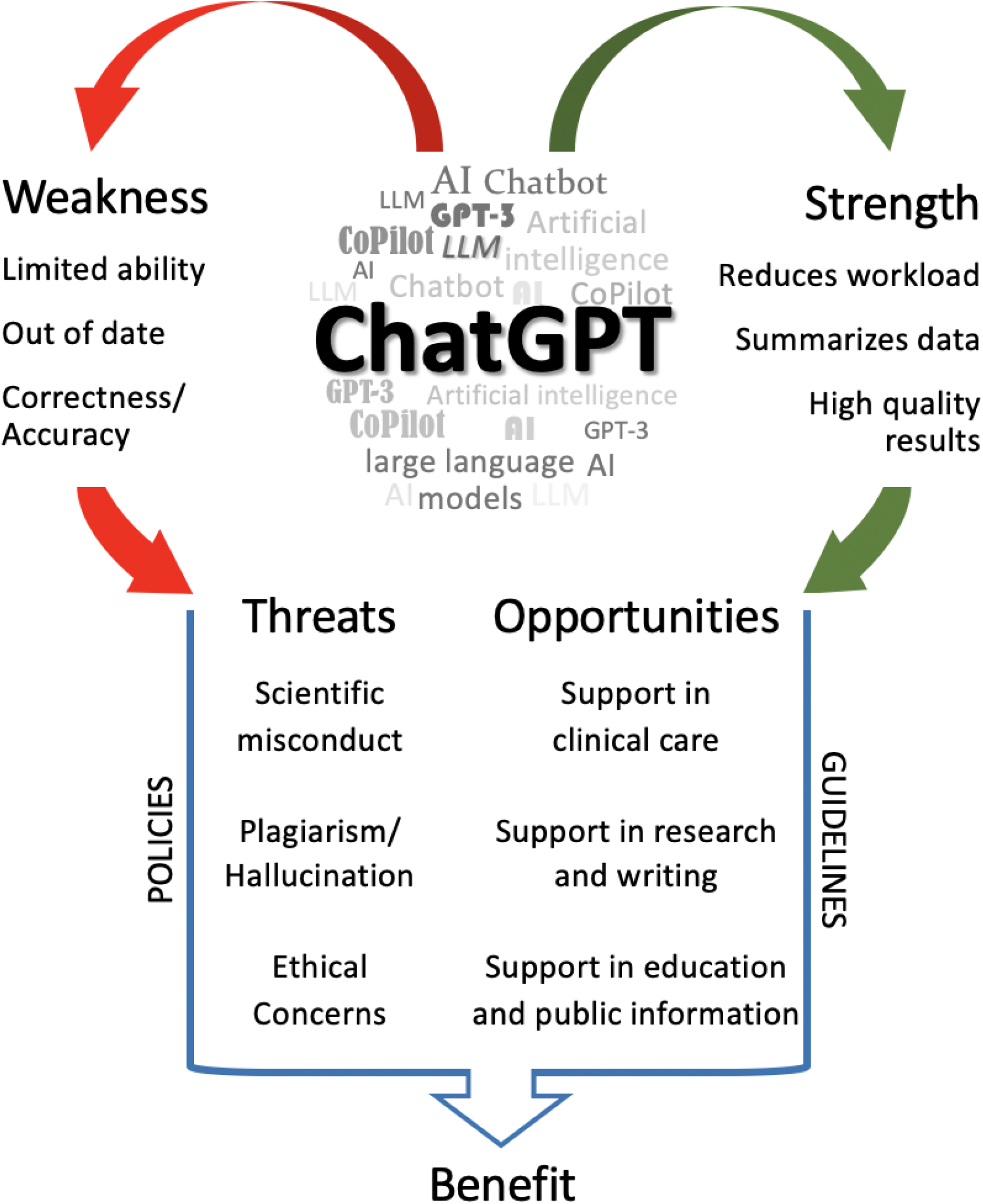
Semi-qualitative SWOT analysis of ChatGPT. Raw data on all information listed above can be found as electronic supplement.

## Discussion

This is the most comprehensive review of ChatGPT to date, summarizing all articles published in PUBMED since its introduction in November 2022 through end of March 2023. In addition to a whole series of metric results, ChatGPT is also critically reviewed in the context of a SWOT analysis. To the best of our knowledge, no similarly comprehensive study on the topic exists to date.

Concerning the article types, it is interesting to see, that so far, no randomized controlled trial has been published about ChatGPT which on the one hand would certainly be difficult to accomplish, but on the other hand is urgently needed. The majority of articles were predominantly of a shorter nature (editorial, letters, features, essays or commentaries).

Journals from the ranks of clinical medicine have published the most articles on ChatGPT, followed by education and others. This resembles the results from authorship. Both aspects, i.e. authorship and journal, show the wide application potential for ChatGPT across many specialist areas as would be expected from a LLM. When considering the impact factor of the journals, it is interesting to see that some articles were published in journals with no impact factor, although even highly reputable fundamental research journals such as Science or Nature as well as clinical journals such as BMJ or Lancet, took up the topic. This proves how important ChatGPT is seen in science, education and clinical work. In the 160 evaluated papers (138 published in Journals with impact factor), there was no significant difference in regard to the impact factor, if the paper was or was not written at least in parts with ChatGPT.

Despite the extensive application possibilities of ChatGPT in many medical, but also non-medical fields, the publication frequency increased rather sluggishly during the observation period, which is somewhat contradictory to the rather spectacular successes of LLM.^1^ Because ChatGPT is also an event of global significance, we deliberately chose the pandemic for comparison, but almost certainly the global health significance was a stronger trigger to address the issue, although no relevant difference was seen during the first four weeks. Interestingly, however, a publication speed, if ascertainable, of 16 days (4-83) was significantly faster than described in other studies on biomedical journals.^31–33^ Beside the spectrum of journals in which ChatGPT publications are made, the proportion of preprint and open access articles could also be decisive in this context. Online publishing has been identified to be strongly associated with reduced submission-to-publication time in multivariate analysis.^33^ Restrictively, it must be mentioned that data on submission speed was only available in about a third of all articles. However, in combination with the higher proportion of quantitative and non-empirical data, we assume all four factors (including open access and preprints) contributed to the fast publication times.

It is difficult to draw a portrait of the authors on this topic due to the distribution pattern as well as the frequency of publication. However, the majority seems to originate from fields of “clinical medicine” which means working with real patients. Education, a frequently mentioned and predestined area of application for ChatGPT, was less present. It should be noted that many authors were not “inexperienced in publishing”, but certainly broke new ground with their publications on the topic. The fact that it seems to be worthwhile to deal with ChatGPT is shown by the average impact factor that could be achieved with a publication on this topic, whereby it was apparently irrelevant whether the article was written with or without the help of ChatGPT. The median impact of 5·144 (with and without the help of ChatGPT) is in a range where only about 12·9% of other journals in a comparison of 13,000 selected scientific journals in 27 major research categories were.^34^ In addition, although it seems tempting to have at least parts of one’s manuscript created with the help of AI, only just under 12% (18/160) made use of it -or at least indicated they had. Clearly recognizing that AI was used for assistance is among the most frequently cited SWOT quotes, but more on that later. So far, however, the use of ChatGPT is not clearly superior or inferior in terms of the impact to be achieved.

Interestingly, ChatGPT was most likely to be identified as having weaknesses in our SWOT analysis, well ahead of strengths, which followed in second place. According to the distribution frequency of the SWOT citations, many authors seem to choose rather a descriptive description of the weaknesses and strengths, but from this much less perspectives or ideas for further handling or further development of Chat GPT developed from their findings. Our assertion is confirmed by the fact, that threats were cited only to 2/3, and opportunities even only to almost 50% in comparison to weaknesses. A SWOT analysis is originally defined as a “*strategic planning and strategic management technique used to help a person or organization identify Strengths, Weaknesses, Opportunities, and Threats related to business competition or project planning*”.^35^ Through a SWOT analysis, favorable and unfavorable internal and external factors achieving the objectives of a venture shall be identified. Some of its advantages include usability and being a “tried-and-true tool” of strategic analysis, points of criticism include limitations like preoccupation leading to neglect, inconsistent compliance with the analysis and domination of certain team members.^36–39^ In order to overcome some of the shortcomings, quotes were analyzed in a modified Delphi process; furthermore, as we intended our SWOT analysis as a starting point for discussion, we considered it as just the right tool for analyzing ChatGPT in its early stages and possibly give some ideas on how to move on from here, particularly in a rapidly changing environment.

So far, ChatGPT has been used to write essays, pass exams, translate knowledge for various peer groups, or write comments on the most diverse topics. During this application it became clear that ChatGPT is apparently “knowledge limited” (until 2021), that source information or even various facts are fictitious, which can only be detected by people with appropriate expertise.

The publications on the topic so far contribute to the fact that, on the one hand, the AI can be improved accordingly, but on the other hand, fields have already been identified in which ChatGPT can presumably be applied safely. These applications include the summarization of large data sets or the quite high linguistic quality of the generated texts.

Overall, caution must be exercised when using ChatGPT, as in several cases sources have been freely invented (hallucination) or copied (plagiarism) and thus the accurateness of the texts created by ChatGPT must always be questioned.

Certainly, no application areas for ChatGPT are, among others, the writing of scientific papers with references, the writing of CVs, or the writing of speeches -in all areas it could already be shown that at least partially completely fictitious passages were formulated by ChatGPT, which did not stand up to a review.

Actually, from our point of view after a concise review, ChatGPT has actually more the state of a toy to explore rather than a reliable tool for scientific working, which doesn’t have to be a bad thing at all, because even through a playful encounter, further strengths could be worked out and further weaknesses could be reflected back to the programmers, which could then be improved. In any case, however, a monopolization is to be avoided, since this could, under certain circumstances, result in major disadvantages such as a ban on viewing source codes or increasing commercialization with ethical-economic imbalances throughout the world. So far, over 30 alternatives for ChatGPT exist including OpenAI playground, Jasper Chat, Bard or Bing AI.^40^ But, in an ideal environment, such large-scale software would always best be open-source.

Another problem of major concern is the ability to detect scientific output by AI. The existing AI Detector software like GPTZero (https://gptzero.me) or related products like GLTR, GPTKit, OpenAI or Output Detector are for example based on scanning for perplexity (rather lower in AI) and burstiness (rather higher in AI).^41^ Their most obvious, clear limitation is that texts are not analyzed for context, but only for writing patterns, which allows the AI to remain undetected. First data on artificial intelligence output detector, plagiarism detector, and blinded human reviewers show promising results: most generated abstracts were detected using the AI output detector, with an AUROC of the AI output detector of 0.94.^42^ Still, further derivatives like GPT-3 or GPT-4 are already “waiting in the wing” - it only remains to hope that test software not only withstands, but hopefully also overtakes the rapid development – always in combination with an alert and suspicious human mind.

How is the phenomenon being dealt with worldwide?

ChatGPT is an AI specialized in written conversations, which makes its application imaginable in almost every area. Its potential has highlighted an absence of any concrete regulation. As by April 2023, ChatGPT is not available in China, nor in various countries with heavy internet censorship like North Korea, Iran and Russia. It is not officially blocked, but OpenAI doesn’t allow users in the country to sign up. Interestingly, several large tech companies in China are developing alternatives.^43^ Italy became the first western country to ban ChatGPT and various other western governments like Germany, Great Britain or Canada are exploring how to regulate AI right now. The U.S. hasn’t yet proposed any formal rules to bring oversight to AI technology.^43^

How is the phenomenon being dealt with in medicine?

During the review, the threat of “remaining undetected” and the associated lack of reproducibility was mentioned.^30^ Suggested solutions include the continuous mention of a possible involvement of ChatGPT,^21^ not as an author,^44,45^ but more as an “acknowledgement”.^46^ This issue has lately been addressed by the world association of medical editors (WAME) clearly stating, that “*AI cannot be an author*” and commenting on responsibility and reproducibility of the human authors.^47^ This is analogously also found in the criteria of the International Committee of Medical Journal Editors (ICMJE).^48^ Major publishers have begun to adopt those recommendations in their own policies.^49^ Other sources recommended inclusion of AI output detectors in the editorial process and clear disclosure if these technologies are used.^42^

Artificial intelligence has always fired human imagination as can be seen from famous movies like Star Trek, Star Wars, Terminator or Aliens - always associated with a resonating, undefined fear that AI may (will?) “overtake” us one day - with potentially deleterious consequences. Despite these easily visualizable and seemingly apocalyptic dangers, one should not condemn the sheer unlimited and fascinating possibilities of artificial intelligence - strict and clear regulation on many levels is necessary to fully leverage the potential. Maybe we should keep a low profile, hence ChatGPT itself points out at least some its weaknesses:

*„As an artificial intelligence language model, I do not have a role in the discussion about ChatGPT in the medical literature. However, I can provide information and answer questions related to my capabilities and limitations as a language model, as well as share insights on how natural language processing technology is being applied in healthcare and medical research*.*”*

## Supporting information

Raw data

## Data Availability

All data produced in the present study are available upon reasonable request to the authors

## Ethics and reporting

All Authors declare that all relevant ethical guidelines have been followed, all necessary IRB and/or ethics committee approvals have been obtained, all necessary patient/participant consent has been obtained and the appropriate institutional forms archived.

## Authors statement

All listed authors declare that all four criteria for authorship in the ICMJE Recommendations are meet individually. All authors confirm that they had full access to all the data in the study and accept responsibility to submit for publication. All authors claim responsibility and accountability for the originality, accuracy, and integrity of the work. Especially, AI and AI-assisted technologies were used twice as examples and clearly marked as such in the manuscript.

## Contributors

All listed authors contributed in the following ways: Literature search (DG, SN, CW, YR, LR, JE, FB, TS), creation of figures (DG, TS), study design (DG, SN, CW, YR, LR, JE, FB, TS), data collection (DG, SN, CW, YR, LR, JE, FB, TS), data analysis (DG, SN, CW, YR, LR, JE, FB, TS), data interpretation (DG, SN, CW, YR, LR, JE, FB, TS), writing (DG, SN, LR, JE, FB, TS).

## Role of the funding source

Institutional funding only; no external funding was utilized for the preparation of the manuscript.

## Declarations of interest

We declare no competing interests.

## Data sharing

Without exception, all data collected for the study, including individual participant data and a data dictionary defining each field in the set, will be made available to others. This also includes additional, related documents will be available with publication and will be made available upon serious request by email from the corresponding author.

## Acknowledgement

The authors would like to thank Dr. rer. nat. Christian Burisch, state of North-Rhine Westphalia, for his help in statistics and the drafting of the manuscript.

## Notes

### Competing Interest Statement

The authors have declared no competing interest.

### Funding Statement

This study did not receive any funding

## References

1. Gilson A, Safranek CW, Huang T, et al. How Does ChatGPT Perform on the United States Medical Licensing Examination? The Implications of Large Language Models for Medical Education and Knowledge Assessment. JMIR Med Educ 2023; 9:e45312. doi: 10.2196/45312

2. Rao A, Pang M, Kim J, et al. Assessing the Utility of ChatGPT Throughout the Entire Clinical Workflow. medRxiv. Preprint posted online 2023 Feb 26. doi: 10.1101/2023.02.21.23285886

3. Benoit JRA. ChatGPT for Clinical Vignette Generation, Revision, and Evaluation. medRxiv. Preprint posted online 2023 Feb 8. doi: 10.1101/2023.02.04.23285478

4. Shorey S, Ang E, Yap J, Ng ED, Lau ST, Chui CK. A Virtual Counseling Application Using Artificial Intelligence for Communication Skills Training in Nursing Education: Development Study. J Med Internet Res 2019; 21(10):e14658. doi: 10.2196/14658

5. Sinhaliz S, Burd L, Du Preez J. How ChatGPT Could Revolutionize Academia - The AI Chatbot Could Enhance Learning, But Also Creates Some Challenges. IEEE Spectrum. 2023 Feb 22. https://spectrum.ieee.org/how-chatgpt-could-revolutionize-academia

6. Roivainen E. I Gave ChatGPT an IQ Test. Here’s What I Discovered. https://www.scientificamerican.com/article/i-gave-chatgpt-an-iq-test-heres-what-i-discovered/, last accessed 01-05-2023

7. Page MJ, McKenzie JE, Bossuyt PM, et al. The PRISMA 2020 statement: an updated guideline for reporting systematic reviews. BMJ 2021; 372:n71. doi: 10.1136/bmj.n71

8. Kambhampati SBS, Vaishya R, Vaish A. Unprecedented surge in publications related to COVID-19 in the first three months of pandemic: A bibliometric analytic report. J Clin Orthop Trauma. 2020; 11(Suppl 3):S304–S306. doi:10.1016/j.jcot.2020.04.030.

9. Wen J, Wang W. The future of ChatGPT in academic research and publishing: A commentary for clinical and translational medicine. Clin Transl Med. 2023 Mar; 13(3):e1207. doi: 10.1002/ctm2.1207.

10. Biswas SS. Potential Use of Chat GPT in Global Warming. Ann Biomed Eng. 2023 Mar 1. doi: 10.1007/s10439-023-03171-8. Epub ahead of print.

11. Shen Y, Heacock L, Elias J, et al. ChatGPT and Other Large Language Models Are Double-edged Swords. Radiology. 2023; 307(2):e230163. doi: 10.1148/radiol.230163. Epub 2023 Jan 26.

12. Zhu JJ, Jiang J, Yang M, Ren ZJ. ChatGPT and Environmental Research. Environ Sci Technol. 2023 Mar 21. doi: 10.1021/acs.est.3c01818. Epub ahead of print.

13. Aubignat M, Diab E. Artificial intelligence and ChatGPT between worst enemy and best friend: The two faces of a revolution and its impact on science and medical schools. Rev Neurol (Paris). 2023; 21:S0035-3787(23)00880-9. doi: 10.1016/j.neurol.2023.03.004. Epub ahead of print.

14. Benichou L; ChatGPT. Rôle de l’utilisation de l’intelligence artificielle ChatGPT dans la rédaction des articles scientifiques médicaux The Role of Using ChatGPT AI in Writing Medical Scientific Articles. J Stomatol Oral Maxillofac Surg. 2023; 101456. doi: 10.1016/j.jormas.2023.101456. Epub ahead of print.

15. Buvat I, Weber W. Nuclear Medicine from a Novel Perspective: Buvat and Weber Talk with OpenAI’s ChatGPT. J Nucl Med. 2023; 64(4):505–507. doi: 10.2967/jnumed.123.265636. Epub 2023 Mar 23.

16. Johnson D, Goodman R, Patrinely J, et al. Assessing the Accuracy and Reliability of AI-Generated Medical Responses: An Evaluation of the Chat-GPT Model. Res Sq [Preprint]. 2023 Feb 28:rs.3.rs-2566942. doi: 10.21203/rs.3.rs-2566942/v1.

17. Yeo YH, Samaan JS, Ng WH, et al. Assessing the performance of ChatGPT in answering questions regarding cirrhosis and hepatocellular carcinoma. Clin Mol Hepatol. 2023 Mar 22. doi: 10.3350/cmh.2023.0089. Epub ahead of print.

18. Rillig MC, Ågerstrand M, Bi M, Gould KA, Sauerland U. Risks and Benefits of Large Language Models for the Environment. Environ Sci Technol. 2023; 57(9):3464–66. doi: 10.1021/acs.est.3c01106. Epub 2023 Feb 23.

19. Gabrielson AT, Odisho AY, Canes D. Harnessing Generative Artificial Intelligence to Improve Efficiency Among Urologists: Welcome ChatGPT. J Urol. 2023; 209(5):827–829. doi: 10.1097/JU.0000000000003383. Epub 2023 Feb 16.

20. Moons P, Van Bulck L. ChatGPT: Can artificial intelligence language models be of value for cardiovascular nurses and allied health professionals. Eur J Cardiovasc Nurs. 2023 Feb 8:zvad022. doi: 10.1093/eurjcn/zvad022. Epub ahead of print.

21. Hosseini M, Horbach SPJM. Fighting reviewer fatigue or amplifying bias? Considerations and recommendations for use of ChatGPT and other Large Language Models in scholarly peer review. Res Sq [Preprint]. 2023 Feb 20:rs.3.rs-2587766. doi: 10.21203/rs.3.rs-2587766/v1.

22. Rao A, Kim J, Kamineni M, Pang M, Lie W, Succi MD. Evaluating ChatGPT as an Adjunct for Radiologic Decision-Making. medRxiv [Preprint]. 2023 Feb 7:2023.02.02.23285399. doi: 10.1101/2023.02.02.23285399.

23. Ali SR, Dobbs TD, Hutchings HA, Whitaker IS. Using ChatGPT to write patient clinic letters. Lancet Digit Health. 2023; 5(4):e179–181. doi: 10.1016/S2589-7500(23)00048-1. Epub 2023 Mar 7.

24. Johnson SB, King AJ, Warner EL, Aneja S, Kann BH, Bylund CL. Using ChatGPT to evaluate cancer myths and misconceptions: artificial intelligence and cancer information. JNCI Cancer Spectr. 2023; 7(2):pkad015. doi: 10.1093/jncics/pkad015.

25. Schorrlepp M, Patzer KH. ChatGPT in der Hausarztpraxis: Die künstliche Intelligenz im Check. MMW Fortschr Med. 2023; 165(6):12–6. German. doi: 10.1007/s15006-023-2473-3.

26. Salvagno M, Taccone FS, Gerli AG. Can artificial intelligence help for scientific writing? Crit Care. 2023 Feb 25;27(1):75. doi: 10.1186/s13054-023-04380-2. Erratum in: Crit Care. 2023; 27(1):99.

27. Zheng H, Zhan H. ChatGPT in Scientific Writing: A Cautionary Tale. Am J Med. 2023 Mar 10:S0002-9343(23)00159-6. doi: 10.1016/j.amjmed.2023.02.011. Epub ahead of print. PMID: 36906169.

28. Baumgartner C. The potential impact of ChatGPT in clinical and translational medicine. Clin Transl Med. 2023; 13(3):e1206. doi: 10.1002/ctm2.1206.

29. Goodman RS, Patrinely JR Jr, Osterman T, Wheless L, Johnson DB. On the cusp: Considering the impact of artificial intelligence language models in healthcare. Med. 2023; 4(3):139–140. doi: 10.1016/j.medj.2023.02.008.

30. Hallsworth JE, Udaondo Z, Pedrós-Alió C, et al. Scientific novelty beyond the experiment. Microb Biotechnol. 2023 Feb 14. doi: 10.1111/1751-7915.14222. Epub ahead of print.

31. Andersen MZ, Fonnes S, Rosenberg J. Time from submission to publication varied widely for biomedical journals: a systematic review. Curr Med Res Opin. 2021; 37(6):985–93. doi: 10.1080/03007995.2021.1905622. Epub 2021 Apr 6.

32. Asaad M, Rajesh A, Banuelos J, Vyas KS, Tran NV. Time from submission to publication in plastic surgery journals: The story of accepted manuscripts. J Plast Reconstr Aesthet Surg. 2020; 73(2):383–90. doi: 10.1016/j.bjps.2019.09.029. Epub 2019 Oct 1.

33. Sebo P, Fournier JP, Maisonneuve H. Is statistician involvement as co-author associated with reduced time to publication of quantitative research in general medical journals? A bibliometric study. Fam Pract. 2019; 36(4):431–36. doi: 10.1093/fampra/cmy115.

34. Joanna W. What’s a Good Impact Factor (Ranking in 27 Categories). https://www.scijournal.org/articles/good-impact-factor, last accessed 22-04-2023

35. SWOT analysis. https://en.wikipedia.org/wiki/SWOT_analysis, last accessed 23-04-2023

36. Dess DG, Lumpkin GT, Eisner AB, McNamara G, eds. The limitations of SWOT analysis. In: Strategic management: text and cases (6th ed.). New York: McGraw-Hill/Irwin. 2012. pp. 82

37. Hill T, Westbrook R. SWOT analysis: it’s time for a product recall. Long Range Planning. 1997;30(1): 46–52. CiteSeerX 10.1.1.469.2246. doi:10.1016/S0024-6301(96)00095-7.

38. Koch JA. SWOT does not need to be recalled: It needs to be enhanced. https://www.westga.edu/~bquest/2000/swot1.html, last accessed 01-05-2023.

39. Chermack TJ Kasshanna BK. The use of and misuse of SWOT analysis and implications for HRD professionals. Human Resource Development International. 2007; 10(4):383–99. doi:10.1080/13678860701718760.

40. Garg S. Top 30 ChatGPT alternatives that will blow your mind in 2023 (free and paid). https://writesonic.com/blog/chatgpt-alternatives/#31-best-chatgpt-alternatives-for-your-to-choose-from, last assessed 23-04-2023

41. Sourceforge. GPTZero alternatives. https://sourceforge.net/software/product/GPTZero/alternatives, last accessed on 23-04-2023

42. Gao CA, Howard FM, Markov NS, Dyer EC, Ramesh S, Luo Y, Pearson AT. Comparing scientific abstracts generated by ChatGPT to original abstracts using an artificial intelligence output detector, plagiarism detector, and blinded human reviewers. bioRxiv; doi: https://doi.org/10.1101/2022.12.23.521610

43. Browne R. Italy became the first Western country to ban ChatGPT. Here’s what other countries are doing. https://www.cnbc.com/2023/04/04/italy-has-banned-chatgpt-heres-what-other-countries-are-doing.html, last accessed 23-04-2023

44. Yeo-Teh NSL, Tang BL. Letter to editor: NLP systems such as ChatGPT cannot be listed as an author because these cannot fulfill widely adopted authorship criteria. Account Res. 2023; 13:1–3. doi: 10.1080/08989621.2023.2177160. Epub ahead of print.

45. Teixeira da Silva JA. Is ChatGPT a valid author? Nurse Educ Pract. 2023; 68:103600. doi: 10.1016/j.nepr.2023.103600. Epub 2023 Mar 7.

46. Siegerink B, Pet LA, Rosendaal FR, Schoones JW. ChatGPT as an author of academic papers is wrong and highlights the concepts of accountability and contributorship. Nurse Educ Pract. 2023; 68:103599. doi: 10.1016/j.nepr.2023.103599. Epub 2023 Mar 4.

47. Zielinski C, Winker M, Aggarwal R, et al. Chatbots, ChatGPT, and Scholarly Manuscripts. WAME Recommendations on ChatGPT and Chatbots in Relation to Scholarly Publications. https://wame.org/page3.php?id=106, last accessed on 23-04-2023.

48. International Committee of Medical Journal Editors MJE. Defining the role of authors and contributors. https://www.icmje.org/recommendations/browse/roles-and-responsibilities/defining-the-role-of-authors-and-contributors.html#two, last accessed 27-04-2023

49. Elsevier. The use of AI and AI-assisted writing technologies in scientific writing. https://www.elsevier.com/about/policies/publishing-ethics/the-use-of-ai-and-ai-assisted-writing-technologies-in-scientific-writing, last accessed 23-04-2023.

